# HPC framework for in-silico trials on 3D virtual human cardiac population to assess drug-induced arrhythmic risk

**DOI:** 10.1101/2021.04.21.21255870

**Authors:** Jazmin Aguado-Sierra, Constantine Butakoff, Renee Brigham, Apollo K. Baron, Guillaume Houzeaux, Jose M. Guerra, Francesc Carreras, David Filgueiras-Rama, Paul A. Iaizzo, Tinen L. Iles, Mariano Vazquez

## Abstract

Cardiotoxicity continues to be a major health issue worldwide due to the imperative need to access new or repurposed drugs that are safe and effective. Accessibility to affordable drugs is also key to ensure access to drugs to all patients who require them. In this work we propose a workflow for an in-silico clinical trial at the 3D biventricular human population level, to assess cardiac pro-arrhythmic risk after administration of a single or a combination of potentially cardiotoxic drugs.

The proposed workflow aims at reproducing gender-specific ionic channel characteristics that determine different responses of patients to drug-induced arrhythmia. To that end a “normal” virtual population of human 3D hearts at rest and exercise/stress (increased heart rate) was analyzed under the influence of drugs, using computer electrophysiology simulations. The changes in ECG, calcium concentration as well as activation patterns on 3D geometry were evaluated for the signs of arrhythmia. Hydroxychloroquine and Azithromycin were used to demonstrate the workflow. Additionally a series of experiments on a reanimated swine heart utilizing Visible Heart^®^ methodologies were performed to verify the arrhythmic behaviour observed in the in-silico trial.

Our results showed similar results to the recently published clinical trials (21% clinical risk vs 21.8% in-silico trial risk). Evidence of transmurally heterogeneous action potential prolongation after a large dose of hydroxychloroquine was an observed mechanism of arrhythmia, both in the in-vitro and the in-silico model. The proposed workflow for the in-silico clinical drug cardiotoxicity trials allows reproducing the complex behavior of cardiac electrophysiology in a population and verifying drug-induced arrhythmic risk in a matter of a few days as compared to the in-vivo trials. Importantly, our results provided evidence of the normal phenotype variants that produce distinct drug-induced arrhythmogenic outcomes.

## Introduction

The fast and safe development of drugs is key to provide access to patients to new, better treatments at lower costs. The same applies to repurposed drugs, particularly when one or more drugs that may have some pro-arrhythmic risk are employed in combination. This issue was stressed during the COVID-19 pandemic with the combined use of potentially pro-arrhythmic drugs as an urgent attempt to repurpose existing treatments to fight the disease [1]. This was the case of the combined use of hydroxychloroquine (HCQ) and/or azithromycin (AZM) [2, 3]. In this context and other clinical situations requiring high-throughput testing, the creation of novel methodologies capable of providing urgent information concerning the cardiotoxic risks of using potentially pro-arrhythmic drugs will provide scientists and physicians a powerful tool in the clinic. Importantly, there are no highly effective clinical predictors, other than QT-interval prolongation and less commonly employed, J-T peak interval; that can provide critical a-priori information regarding the potential risks for certain patients with normal QTc intervals to develop QT-prolongation after the administration of one or various drugs that may have cardio-toxic side-effects.

The function of various cardiac ion channels can be significantly modified by environmental conditions (i.e. hormones, electrolyte concentrations and pH), which can in turn have substantial effects on the overall electrical profile. Males and females present different risks for drug-induced arrhythmias and QT-interval prolongation due to sex-specific hormones [4, 5]. In the case of COVID-19, hypokalemia has been identified as a prevalent condition in patients, which may increase further the risk of QT prolongation [6]. Further, the combined administration of several drugs increases the complexities of understanding the associated clinical implications, especially given a very limited information about drug interactions.

Computational methods have been an important component for the study of drug-induced arrhythmias [7–11]. However most of the current studies focus on single cell population analyses (i.e. [12, 13]) or full 3D biventricular anatomies analyzing a variety of doses or a few anatomies (i.e. [14–16]). A human population described using a highly detailed, gender-specific human anatomy has never been reported. The use of full biventricular anatomies is crucial to assess drug effects on the full heart and understand the mechanisms of the arrhythmic risk beyond a single cell behaviour. Furthermore, the use of the full heart anatomy allows including other risk factors like electrolyte imbalances, myocardial infarction, ischemia, or cell heterogeneity.

The workflow, proposed in this paper starts by designing a “normal” virtual population of human 3D hearts. These hearts are then subjected to the effects of the drugs that are potentially cardiotoxic. Subsequently, computer simulations are carried out in normal and stressed conditions (increased heart rate) and the changes in ECG, calcium concentration as well as activation patterns on 3D geometry are evaluated for the signs of arrhythmia. To model a “normal” population, we defined a set of hypotheses. One requirement towards the creation of a virtual population was to include gender-specific ion channel phenotypes; in order to reproduce the gender-specific risks to drug QTc prolongation. Another requirement was to create a diverse ion channel description. We hypothesize that the diversity of experimentally calibrated cardiomyocyte ion channel conductances may resemble the variabilities of ion channel expressions observed clinically [17, 18]. We further hypothesize that these same variabilities may apply to both male and female genders. These hypotheses were employed given that there is no experimental information regarding normal inter-subject ion channel variability. The main contributions of this paper are:

- A methodology to define a “normal” virtual population of 3D hearts for in-silico cardiotoxicity trials using diverse ion channel phenotypic description and including gender-specific ion channel phenotypes in order to reproduce gender-specific risks to drug QTc prolongation [11, 17–19]
- The use of published IC50 and drug effective plasma concentrations from a variety of available sources at the time of the COVID19 pandemic. The methodology for the administration of drugs followed Beattie et al [10], with the combination of the two drugs assumed to have additive effect, as in Delaunois et al [9]
- Develop a reliable methodology to identify the risks of cardiotoxicity in-silico using virtual stress test and classify the responses within the virtual population to the administered drugs. This arrhythmic risk evaluation is based on clinical case reports of exercise-induced arrhythmias related to the administration of antimalarial drugs [20].

As a result we show that it is possible to assess the pro-arrhythmic risk produced by the administration of one or more potentially arrhythmic drugs using the proposed methodology. The generated population can reproduce the arrhythmic risk observed clinically after the administration of HCQ and AZM. Published effective plasma concentrations and IC50 values of HCQ and AZM employed to administer a variety of doses to the human virtual population provided meaningful information regarding clinically observed pro-arrhythmic risk. Finally, we employed reanimated swine hearts to observe acute arrhythmic features induced by the administrations of high doses of drugs and compare the observed arrhythmic mechanisms to the ones observed in our virtual population.

The contributions of this paper represent a gigantic leap from existing experimental single cardiomyocyte data to full 3D cardiac physiologic responses that reproduce the behaviour of a human population, which open the possibilities of full organ-scale, translational, in-silico clinical trials to assess drug-induced arrhythmic risk.

## Materials and methods

The detailed biventricular anatomy, of a female (24yo, BMI 31.2) ex-vivo, perfusion-fixed human heart, was segmented from high-resolution magnetic resonance imaging (MRI) data. This heart was obtained from an organ donor whose heart was deemed not viable for transplant, via the procurement organization, LifeSource (Minneapolis, MN, USA). All donors had consented to donating their organs for transplant prior to death. This specimen remains a part of the Visible Heart^®^ Laboratories’ Human Heart Library at the University of Minnesota. The uses of heart specimens for research and the laboratory was appropriately consented and signed by the donor families and witnessed by LifeSource, and the research protocol was reviewed by the University of Minnesota’s Institutional Review Board and LifeSource’s Research Committee. All data was deindentified from its source and it is medically exempt from IRB. The heart was cannulated and perfusion-fixed under a pressure of 40 - 50 mmHg, with 10% phosphate buffered formalin; which preserved it in an end-diastolic state. High resolution images of this specimen were acquired via a 3T Siemens scanner: with 0.44 times 0.44 mm in-plane resolution and slice thickness of 1 to 1.7 mm, which provides detailed endocardial trabeculae and false tendons with ≈ 1*mm*^2^ cross sectional area.

A volumetric finite element mesh (of 65.5 million tetrahedral elements) was created [21], with a regular element side length of 328 micron using ANSA (BETA CAE Systems USA Inc.), with a rule based fiber model [22] and transmural cell heterogeneity (endocardial, midmyocardial and epicardial). The finite element model of electrophysiology was solved using Alya, a source available high performance computing software developed at the Barcelona Supercomputing Center, employing the monodomain approximation to the anisotropic electrical propagation [23, 24]. This software is designed from scratch to run efficiently in high performance computers, with a tested scalability up to cores [25–29]. The monodomain approximation to the solution of electrophysiology was programmed within Alya. Briefly, the equations for electrical propagation and the system to be solved is[TODO:REF]:

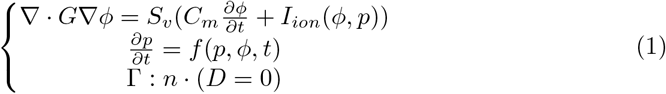

where *C*_*m*_ refers to the membrane capacitance; *ϕ* is the transmembrane potential, *G* is the conductivity tensor which defines the anisotropic conductivity in the cardiac muscle; *S*_*v*_ is the surface to volume ratio of the cardiomyocytes; and *I*_*ion*_ is the solution of the ODEs that define the cardiomyocyte ion channel model based on the one published by O’Hara-Rudy [30]; *f* is this system of ODEs, which is dependent on the transmembrane potential *ϕ* and a set of parameters p. Zero flux is defined at the boundary Γ of the domain. A Yanenko operator splitting technique was applied to discretize the system. The ODEs system is solved using a forward Euler explicit scheme. The PDE is solved using a Crank-Nicolson scheme. The problem domain is partitioned using METIS [31] into several sub-domains and parallelized using MPI.

The fast endocardial activation due to the Purkinje network was approximated by defining a fast endocardial layer with an approximate thickness of 348 microns. The anisotropic diffusion coefficient of both, the purkinje layer (10x myocardial diffusion) and the myocardial tissue were obtained after thorough analysis of the total activation times, QRS duration and epicardial conduction velocities to obtain normal clinical values (diffusion of 3.7*cm*^2^*/s* in the fiber direction and 1.2*cm*^2^*/s* in the transverse directions) [21]. The convergence on conduction velocity for the electrophysiology setup can be found in the supplemental data. Pseudo-electrocardiograms (pseudo-ECGs) were calculated by positioning each biventricular model within a generic torso (constructed from computed tomography scan of a patient) and recording the cardiac potential (integrals over the spatial gradients of the transmembrane voltages within the biventricular cardiac tissue) at the approximate locations of the right arm (RA), left arm (LA) and left leg (LL).

The transmural myocyte heterogeneity was added by processing the solutions of the heat equation obtained during fiber generation [22]. Given two solutions Φ_*tm*_ and Φ_*epi*_ (Φ_*tm*_ obtained by applying temperatures 0 on epicardium, 1 on RV endocardium, −2 on LV endocardium; Φ_*epi*_ by applying 1 on epicardium and 0 on LV and RV endocardium). The celltype was assigned using the following rule (c.f. Fig. 1):

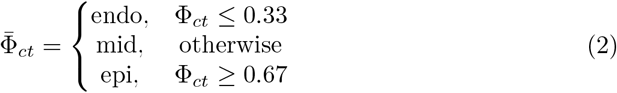

where

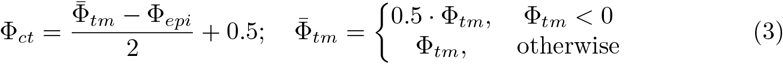

**Fig 1.**
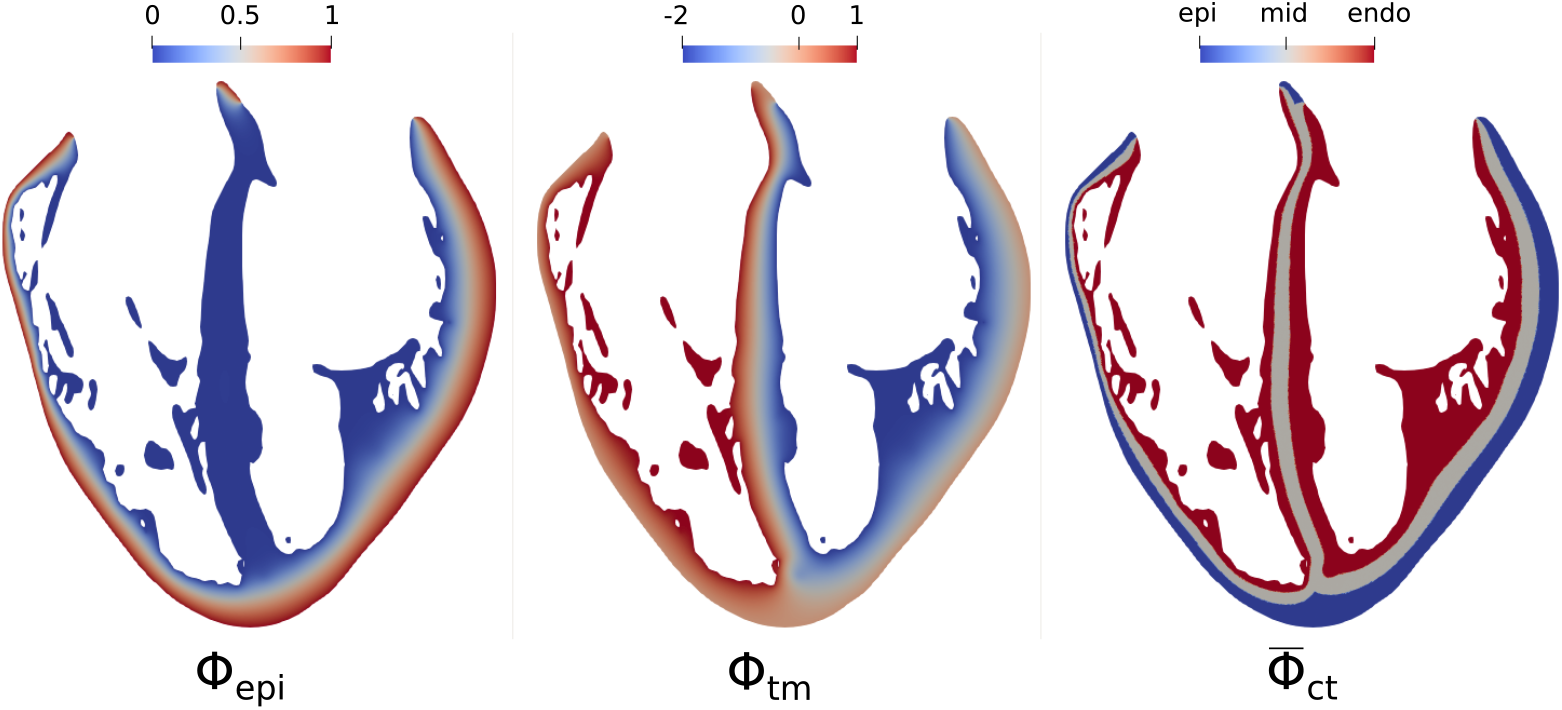
Cell type definition. 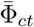 calculated from the heat equation solutions Φ_*tm*_ and Φ_*epi*_. Shown is approximately long axis 4 chamber view, RV on the left and LV on the right.

The transmural myocyte heterogeneity was applied following the modifications to the endocardial cell type published by O’Hara and Rudy [30]. The locations of the activation regions within the biventricular cavities was set following the work of Durrer [32]. The initial activation regions (IARs) were: 1-2. two activation areas on the RV wall, near the insertion of the anterior papillary muscle (A-PM) 3. high anterior para-septal LV area below the mitral valve 4. central area on the left surface of the septum 5. posterior LV para-septal area at 1*/*3 of the apex-base distance; as shown in Fig. 2. The activation magnitude was −80mV applied for 5ms at each of the specified locations, reproducing a spherical stimulus of 0.2cm radius at the required heart rate. Pseudo-electrocardiograms (pseudo-ECGs) were calculated by positioning each biventricular model within a generic torso, as can be observed in the Fig. 2 following the methodology published by Gima and Rudy [33]. The electrical propagation throughout the torso was not computed.

**Fig 2.**
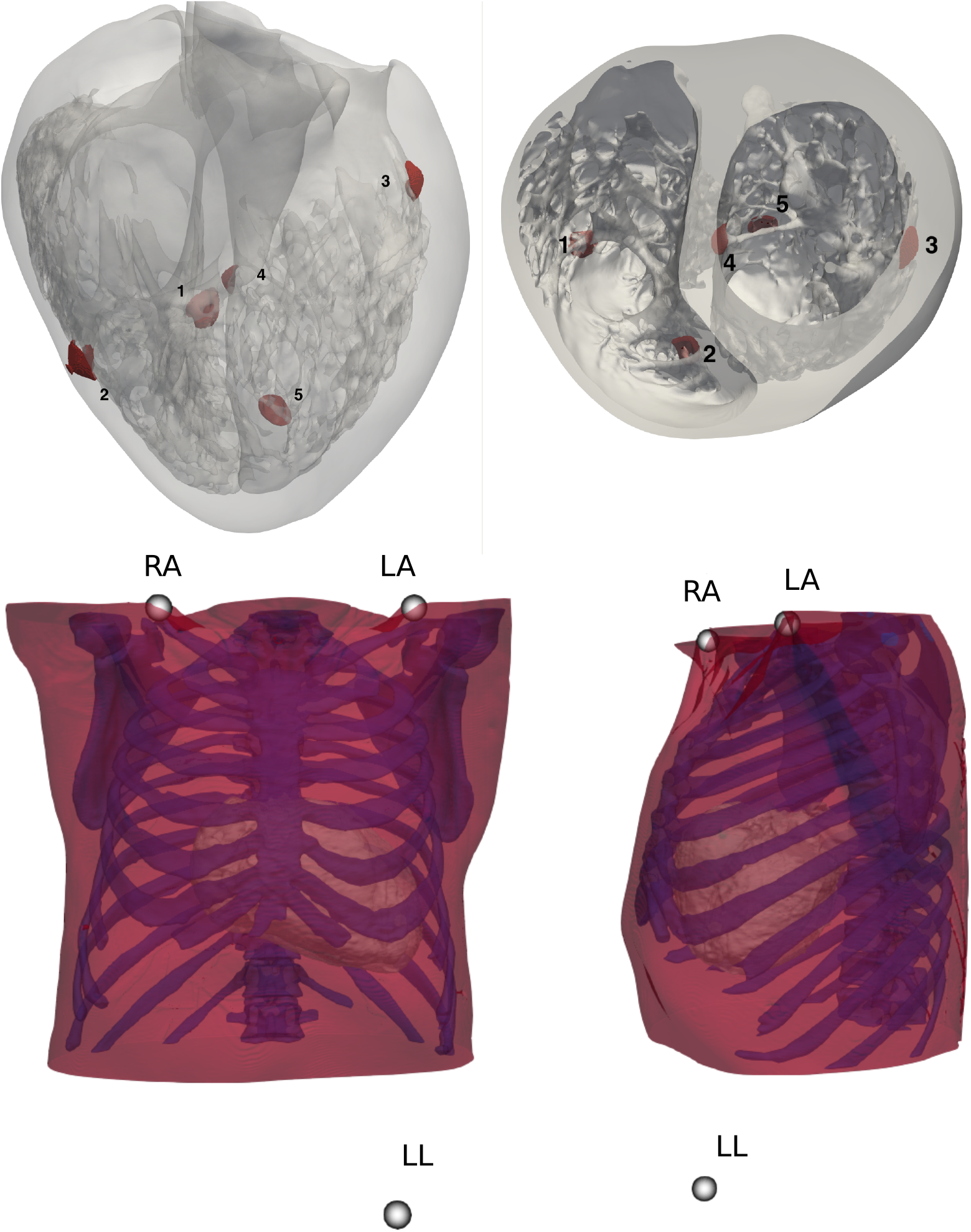
First row shows the high resolution human biventricular heart anatomy employed. The initial stimuli locations are shown in red. The activation magnitude was -80 mV applied for 5 ms, reproducing a spherical stimulus of 0.2 cm radius. Below, the torso employed as reference to spatially locate the pseudo-ECG leads. The heart is shown registered within the torso.

The O’Hara-Rudy [30] human cardiac ionic model with the conductance modifications to the sodium channel suggested by Dutta [34], as used by the FDA as the basis for the CiPA initiative for proarrhythmic risk assessment, was employed to compute the cardiomyocyte action potentials. To study the gender-specific cardiotoxicities within a human heart population, all the changes to ion channels sub-unit expressions listed by Yang et al. [19], were applied to generate both male and female phenotypic cohorts. The spectrum of phenotypes within the normal population was set up employing the experimentally calibrated normal population data published by Muszkiewicz et al. [18]. Five ion channels that have the highest influence on action potential durations were selected: INa, IKr, IKs, ICaL and INaL. The first and third quartiles of the above channel conductances (see Table 1) were sampled in a combinatorial manner in order to capture the bounding values of ion channel function. Therefore, the problem was reduced to 2^5^ permutations (1st and 3rd quartiles) for the 5 channels, for a total of 64 subjects: 32 unique male and 32 female phenotypes. Additionally to the normal baseline population, Hypokalemic population was obtained by reducing the extracellular potassium concentration to 3.2 mmol/L in the normal population, as an example of a population that would potentially be affected even more by the drugs.

**Table 1.** Scaling factors by which each ion channel is modified in a combinatorial manner.

| Quartile | GKr | GKs | GNaL | GNa | GCaL |
| --- | --- | --- | --- | --- | --- |
| Q1 | 0.80 | 0.75 | 0.75 | 0.80 | 0.75 |
| Q3 | 1.25 | 1.25 | 1.25 | 1.20 | 1.25 |

Hydroxychloroquine (HCQ) and Azithromycin (AZM) were used to illustrate the proposed framework. The effects of the drugs affecting the ion channel conductances were incorporated following the methodology of Mirams et al. [35] using a multi-channel conductance-block formulation for each drug assessed. Peak plasma concentration at clinical oral doses and IC50 values for AZM [36], peak plasma concentrations for HQM [37] and its IC50 [38] were: 800 and 400 mg single oral dose of HCQ and combined administration of 200 and 400 mg HCQ and 500 mg AZM. All drug-related concentrations and IC50 values employed in this study can be found in Tables 2 and 3. The assumptions regarding the interactions of both drugs were based on the additive effects of each drug on the affected ion channels described by the following equations (with *D*_*i*_, IC50_*i*_ and *h*_*i*_ being the concentration, IC50 and h values for the *i*-th drug and *g,g*^*′*^ – the channel conductance before and after the application of drugs):

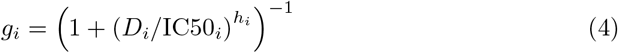

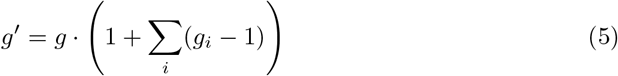

**Table 2.** Plasma concentrations employed for each oral dose tested (obtained from [18, 38]).

| Oral Doses | HCQ 200 mg | HCQ 400 mg | HCQ 800 mg | AZM 500 (3-day) |
| --- | --- | --- | --- | --- |
| Peak Plasma Concentration ($\mu$ mol/lit) | 1.19 | 2.97 | 4.7 | 0.59 |

**Table 3.** IC50 values for each channel affected by HCQ and AZM. Data obtained from [37, 38].

| Drug | IKr IC50 (μmol) | ICaL IC50 (μmol) | IKs IC50 (μmol) | INa IC50 (μmol) | IK1 IC50 (μmol) | INaL IC50 (μmol) |
| --- | --- | --- | --- | --- | --- | --- |
| HCQ | 5.57 | 22 | NA | NA | NA | NA |
| AZM(chronic effect) | 219 | 66.5 | 184 | 53.3 | 43.8 | 62.2 |

The ion channel conductance variations and the drug blocking effects on the ion channel models were solved in zero-dimensional (0D) models (endocardial, midmyocardial and epicardial) until steady state of the calcium transient was achieved (root mean square error, RMSE, between consecutive beats smaller than 1e-7(*µmol*) for 3 consecutive beats). The results of the 0D models provide initial conditions to parameterize the 3D finite element simulations. Simulations on the 3D mesh of 3 to 5 beats were solved to achieve steady state at two different basic cycle lengths (BCL): 600 (Baseline) and 400 ms (Stress). A total of 896 simulations were run, each one using 640 cores on the Joliot-Curie Rome supercomputer(GENCI, CEA, France). An approximate total of 2.3 million core hours were employed.

The QTc and QRS values were measured using the three pseudo-ECG leads. An example pseudo-ECG of a male and female subject is shown in Figure 3, including an example of the calculation of the QRS and QT markers. It is important to state that at this point of our work, we did not aim to validate an electrocardiogram calculation. Instead, the pseudo-electrocardiogram was employed to obtain markers at baseline and after the administration of drugs to quantify their effect. Both Framingham and Bazett formulae were used to calculate the QTc from the cohort with a BCL of 600 ms. A surrogate marker for contractility was obtained by calculating the integral of the calcium transient throughout the anatomy. The magnitude of the peak Calcium, the time to peak Calcium, Calcium 90 and the magnitude of the T-wave were also quantified in each simulation. Data were analysed using RStudio and Orange [39]. The relative importances of each phenotype on each marker was assessed using the LMG (Lindeman, Merenda, and Gold’s (LMG) method [40]), implemented in the relaimpo R package [41]. The risk characterization was used to identify the currents that have an individual contribution to a multiple regression model. The entire dataset of simulations which were characterised as having risk (including all drug administrations) were used. Two response variables were employed: QTc interval value and QTc-interval prolongation. The multiple linear regression was performed with the ion channel conductance factors as regressors to either of the response variables.

**Fig 3.**
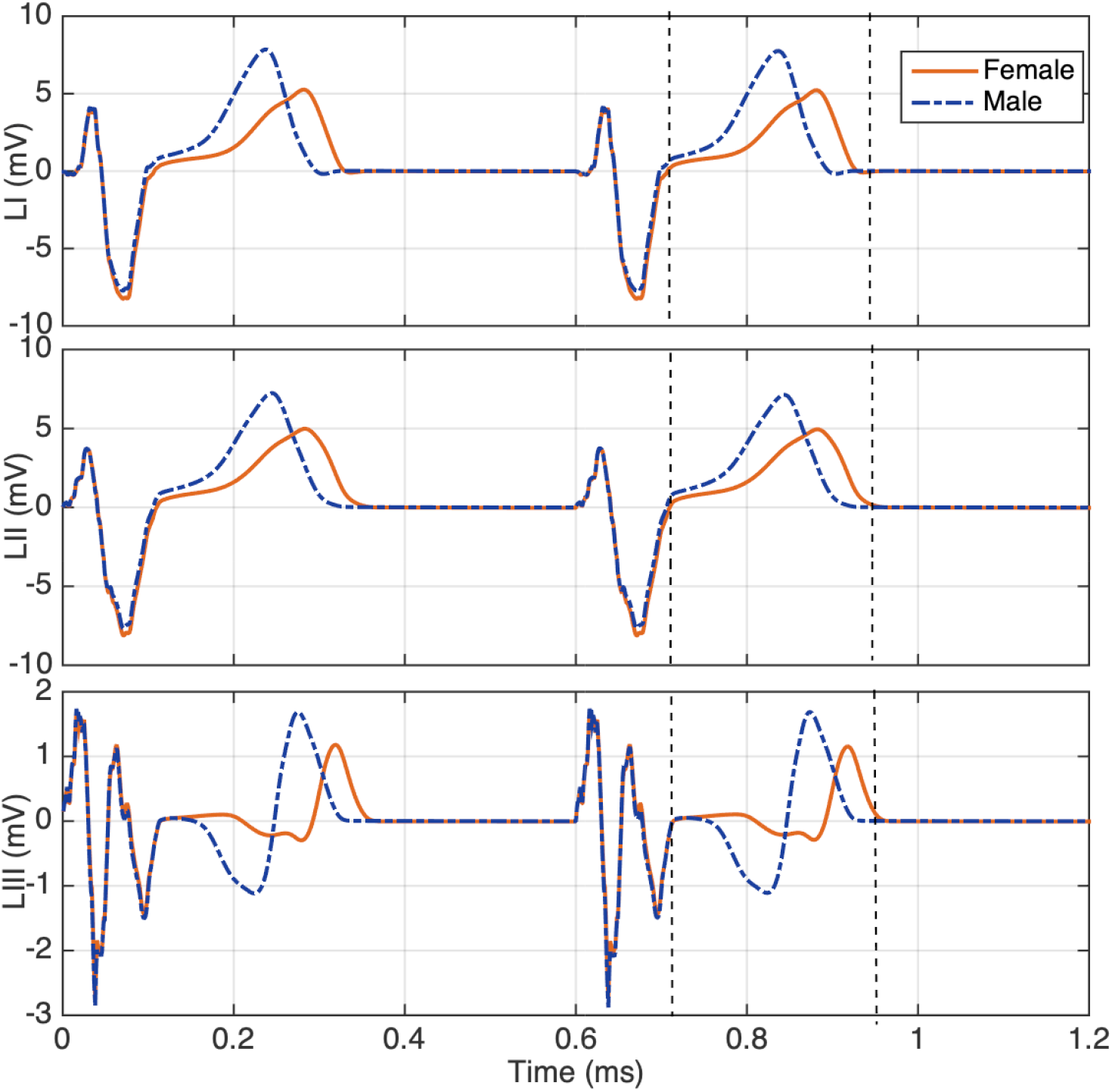
Pseudo-electrocardiogram calculated at three standard lead locations: LI, LII and LIII of a male and female subject of our population. QRS and QT intervals as calculated for our analysis are marked within each lead. Both markers were determined using the three leads.

The stress test to assess arrhythmic risk of each subject was performed by running the population at the BCL of 400 ms (150 bpm). A subject at risk was defined by observing spontaneous arrhythmic or dangerous rhythms, which included left bundle branch block, ventricular tachycardia or QT-interval greater than 390 ms at a 400 ms bcl, which lead to observable asynchronous activation, result of the tissue remaining in the refractory period. Figure 4 shows examples of the classifications performed on the pseudo-ecg data after the stress test. A semi-automatic algorithm was employed to estimate the QT interval duration of all the pseudo-ecg database. Detailed analysis was also performed by observing the results of the electrophysiological propagation in the human heart of interest.

**Fig 4.**
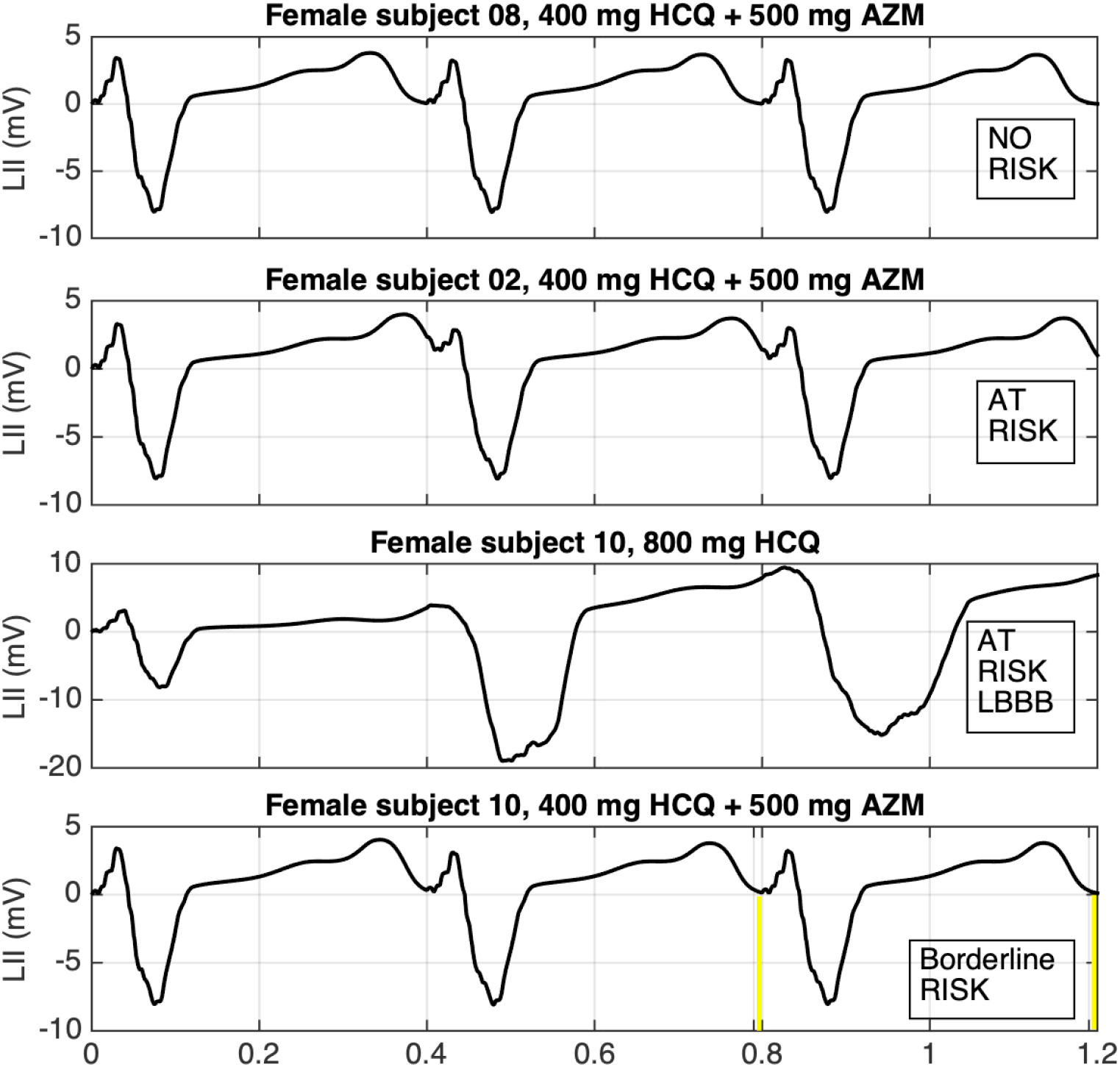
Analysis of the Pseudo-ECG to characterise risk after the stress test. Pseudo-electrocardiograms resulting from the stress test employed to characterize risk. Four different cases were selected. Risk is characterised as the incomplete repolarization of the heart before the next heart beat occurs. When end of repolarization and activation occurs at the same time (marked by yellow vertical bars), the subsequent beat presents lengthening of the QRS due to dyssynchronous activation produced in regions where the AP is still in a refractory phase. The 10 ms window between the 390 and 400 ms of the last two cardiac cycles is highlighted in yellow.

### In-vitro Experimentation on Reanimated Swine Hearts

To assess the arrhythmic behaviour of the computational models with regards to electrophysiological and functional effects in response to HCQ administration, an experimental in-vitro setup employing reanimated swine hearts was used. All animals received humane care in compliance with the ‘Principles of Laboratory Animal Care’, formulated by the National Society for Medical Research, and The Guide for the Care of Laboratory Animals, published by the National Institutes of Health. This research protocol was ethically approved by the University of Minnesota Institutional Animal Care and Use Committee. The IACUC protocol number is 2006A38201 “Swine Isolated Heart Model”. The precise experimental protocol was published previously [42]. Electrical potentials and mechanical function were monitored throughout the experiment. These supplemental experiments were performed on a reanimated swine heart utilizing Visible Heart^®^ methodologies. Briefly, swine hearts were continuously perfused with a clear, modified Krebs-Henseleit buffer according to previously described methodologies [43, 44]. After providing a single 30 J shock, each heart elicited and sustained an intrinsic rhythm and associated hemodynamic function (more details in supplement).

#### Mechanical and Electrical Data Acquisition

Pressure volume loops were measured using clinically available conductance catheters and a data acquisition system (CD Leycom, Netherlands). Conductance catheters were placed in both left and right ventricles and calibrated to the cardiac function, the data was sampled at a rate of 250Hz. Local Monophasic Action Potentials (MAPS) were recorded from both ventricles with MAP4 catheters (Medtronic, Minneapolis, MN, USA) positioned in the RV and LV endocardium and LV anterior epicardium, and the rise time and time to decay were measured for 10 consecutive MAP waveforms for each treatment group. Additionally, 2 decapolar catheters were sutured onto the epicardial surfaces of both ventricles to record the relative changes in electrical activations along the ventricles; in order to capture evidence of dispersion of repolarization. 3-lead surface electrocardiograms were recorded throughout the duration of the study to measure global electrical activity as shown in Figure5.

**Fig 5.**
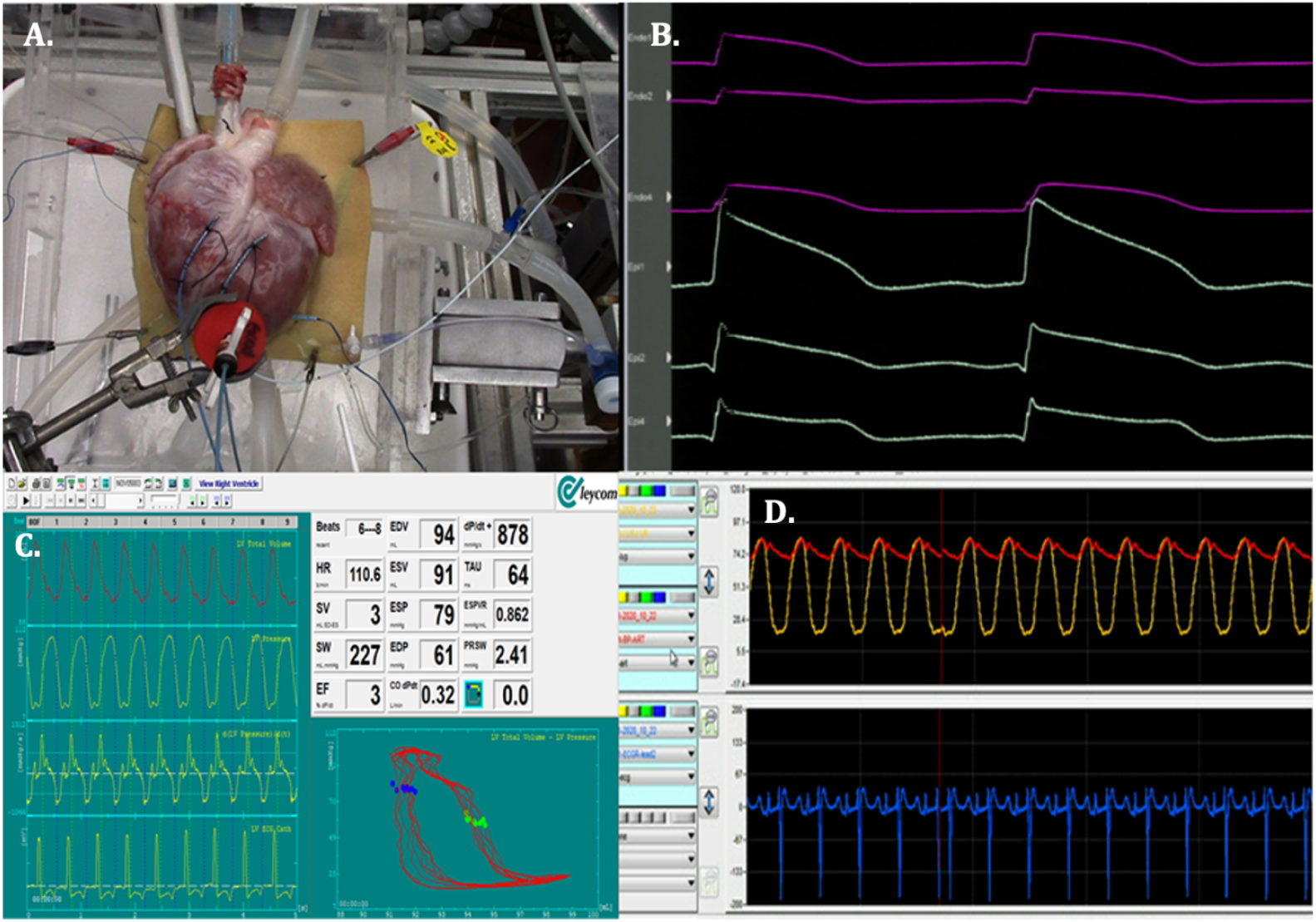
A) An external view of the reanimated swine heart. Two decapolar catheters were sutured to the epicardial surfaces of both the left and right ventricles and a custom device holds the MAPS4 catheter in constant contact with the LV epicardium. B) Electrical activations in LV and RV as measured from decapolar catheters. C) PV loops are measured from conductance catheters placed within both the LV and RV. D) Arterial (red trace) and LV pressures (yellow trace) are recorded along with the 3-lead surface ECG (blue trace)

The datasets were collected at four separate time points: baseline, during drug infusion of 25 mg of HCQ, after a 10 minute washout period (in which buffer was replaced twice), during a second drug infusion of 25 mg of HCQ and then a final 50 mg infusion of HCQ after another 10 minutes washout period. The drug was prepared by dissolving 25 mg of HCQ in 10L of recirculated buffer (equivalent to 7.44 µmol/lt) and allowed to equilibrate for 2 minutes before data collection. Doses were experimentally derived to observe the greatest function differential from controls to treatment with HCQ.

## Results

### In-silico experiments

Median, interquartile range, mean and standard deviation of all markers measured are shown in Table S1. Mean and standard deviation is included in order to compare the results to clinically published data.

The QTcFra histogram of the simulated population classified by gender at baseline, hypokalemia and after the administration of a variety of oral doses and combined administration of HCQ and AZM are shown in Fig. 6. The QTc prolongation in the entire cohort is shown in Fig. 7.

**Fig 6.**
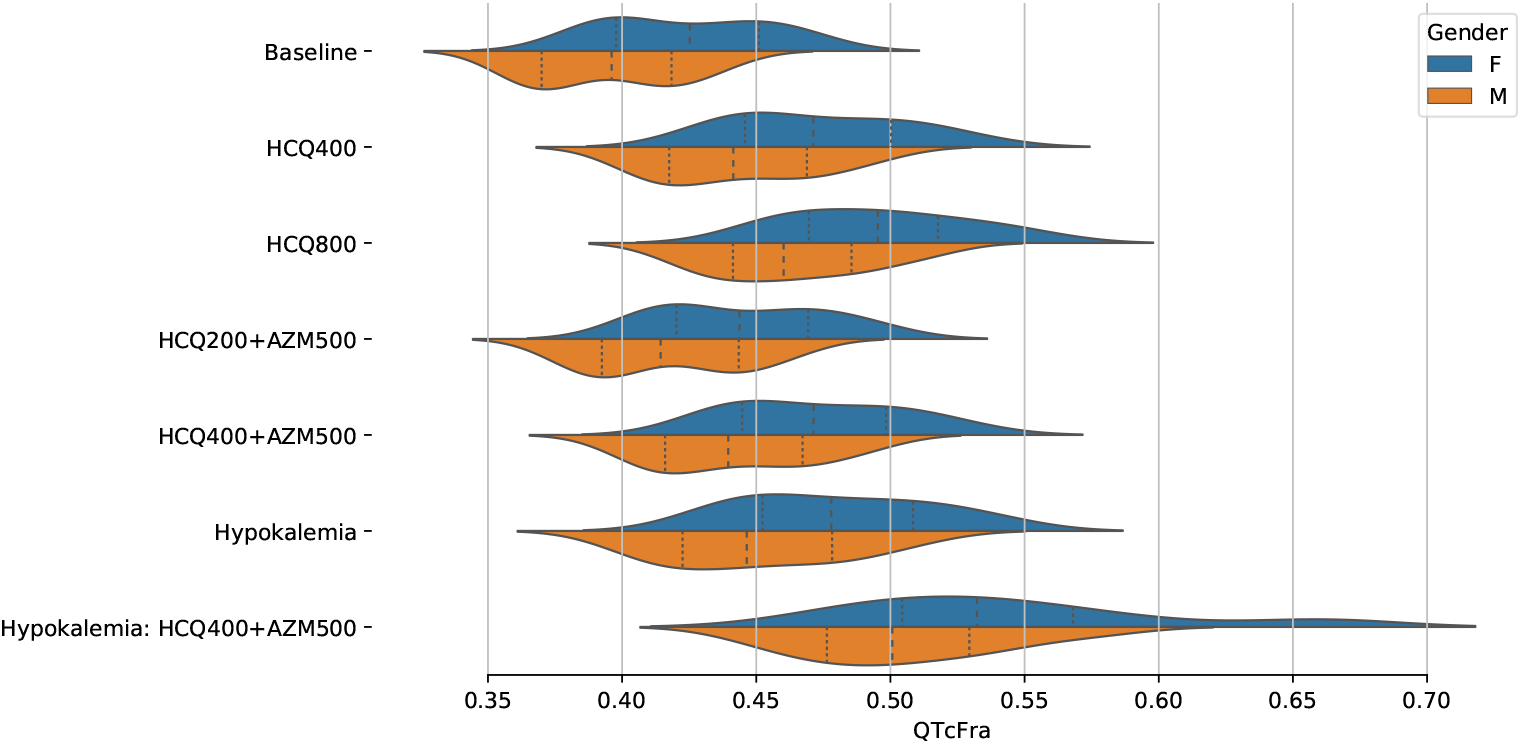
**Histogram of the QTcFra values** of the entire population classified by gender during baseline and after the administrations of 400 and 800 mg HCQ and the use of 200 and 400 mg HCQ in combination with 500 mg of AZM under normal conditions and under hypokalemia (3.2 mmol/L). QTcFra corresponds to the baseline BCL of 600ms.

**Fig 7.**
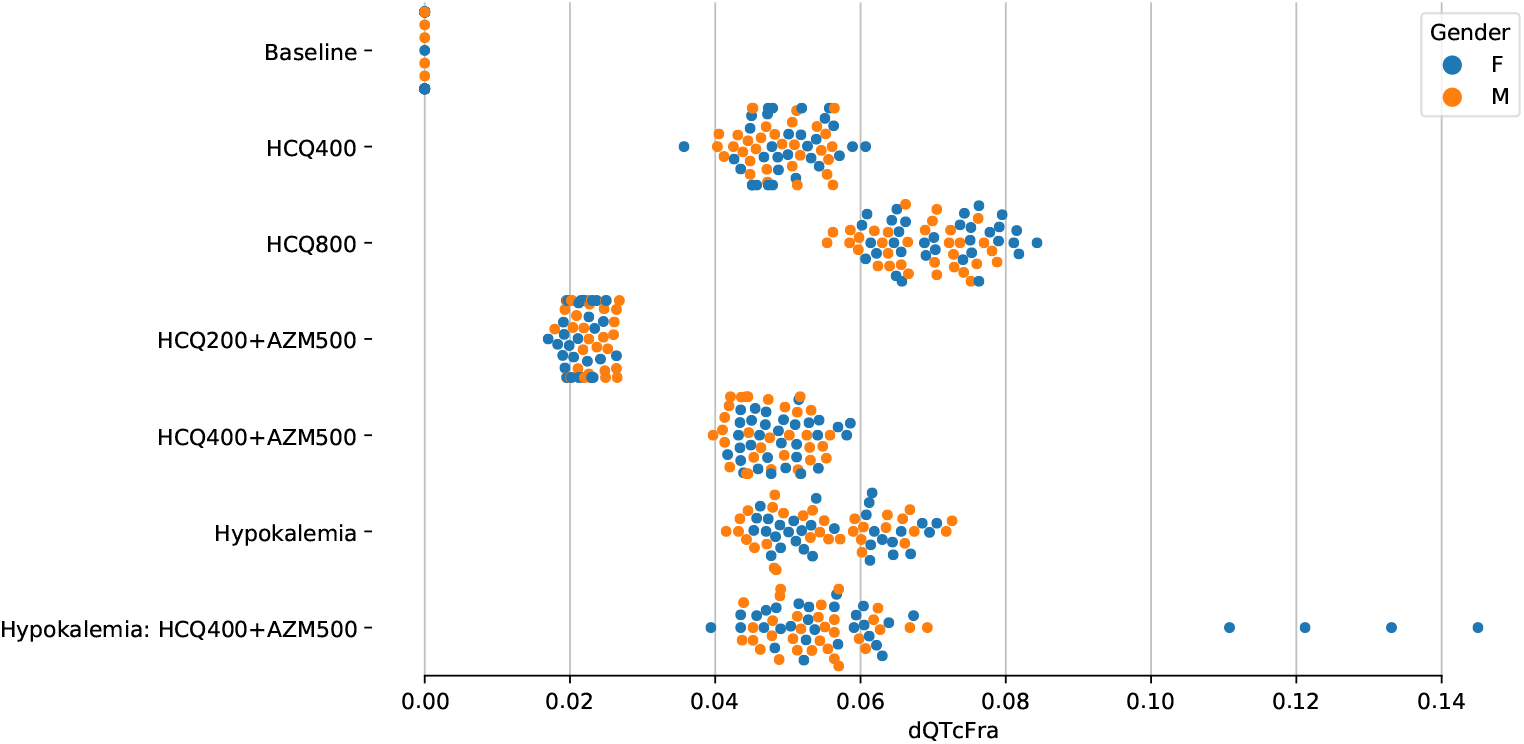
QTcFra (s) prolongation classified by gender (dQTcFra). Both genders provide approximately similar QTcFra prolongations. Four female subjects with Hypokalemia and HCQ400 + AZM500 presented extreme prolongation that lead to ventricular tachycardia. The QTcFra corresponds to the baseline BCL of 600 ms.

A preliminary classification of the virtual clinical trials based on QTc values (at BCL = 600 ms) is shown in Table 4. There were no arrhythmic events in any of the subjects at this BCL. The number of males and females with QTc Framingham values higher than 500 ms are included. It is noticeable that females present a higher percentage of the population with increased QTc-interval values and risk.

**Table 4.** Classification based on QTc Framingham values from the virtual clinical trial, corresponding to BCL of 600 ms.

| Human In-silico Trial | Subjects with QTcFra > 500 (%) | Female N = 32 | Male N = 32 |
| --- | --- | --- | --- |
| Baseline | 0 | 0 | 0 |
| HCQ 400 mg | 9, (14%) | 8 | 1 |
| HCQ 800 mg | 17, (26%) | 22 | 4 |
| HCQ 200 mg + AZM 500 mg | 1, (1%) | 1 | 0 |
| HCQ 400 mg + AZM 500 mg | 7, (10%) | 7 | 0 |
| Hypokalemia (Ko=3.2 mmol) | 14, (21%) | 11 | 3 |
| Hypokalemia (Ko=3.2 mmol) + HCQ 400 mg + AZM 500 mg | 41, (64%) | 24 | 17 |

The solutions for the full cohort after a stress test showed numerous spontaneous arrhythmic responses that were employed to characterize the risks of drug-induced arrhythmia. For example, LBBB was observed in two cases after the administration of 800 mg of HCQ; one case after the administration of 400 mg of HCQ and 500 mg of AZM; and four at a hypokalemic state after the administrtion of HCQ 400 mg and AZM 500 mg. Four cases of ventricular tachycardia were observed at a hypokalemic state after the administration of HCQ 400 mg and AZM 500 mg. The virtual patients at risk were classified according to the QTcFra prolongation as described in Fig. 4 after the administration of the drugs. Results are shown in Fig. 8. A classification based on risk, including gender differences of risk are shown in Table 5. It is important to note that the virtual stress test was able to reproduce the complexity of drug-induced arrhythmia, where QTc was not a definite marker of arrhythmic risk, as it is shown in detail in Fig. 9. The QTc value of each virtual patient at baseline, hypokalaemia, and the progression of the QTc prolongation after the administration of one or more drugs are plotted. To the right, the QTc prolongation in subjects that present no arrhythmic response, and to the left, the subjects that present an arrhythmic risk after being classified by the stress test for each drug intervention. It is also clear by the slope of the drug-induced QTc prolongation, the effect of each drug regime. Fig. 9 clearly shows that QTcFra prolongation values do not fully characterise the drug-induced arrhythmia within the virtual population, as is known to be the case in the clinical setting. This precise observation makes the stratification of patients that can develop drug-induced arrhythmias a major unsolved problem clinically. It was observable that patients with the same QTcFra at baseline could present two distinct responses to drug administration during the stress test.

**Table 5.**
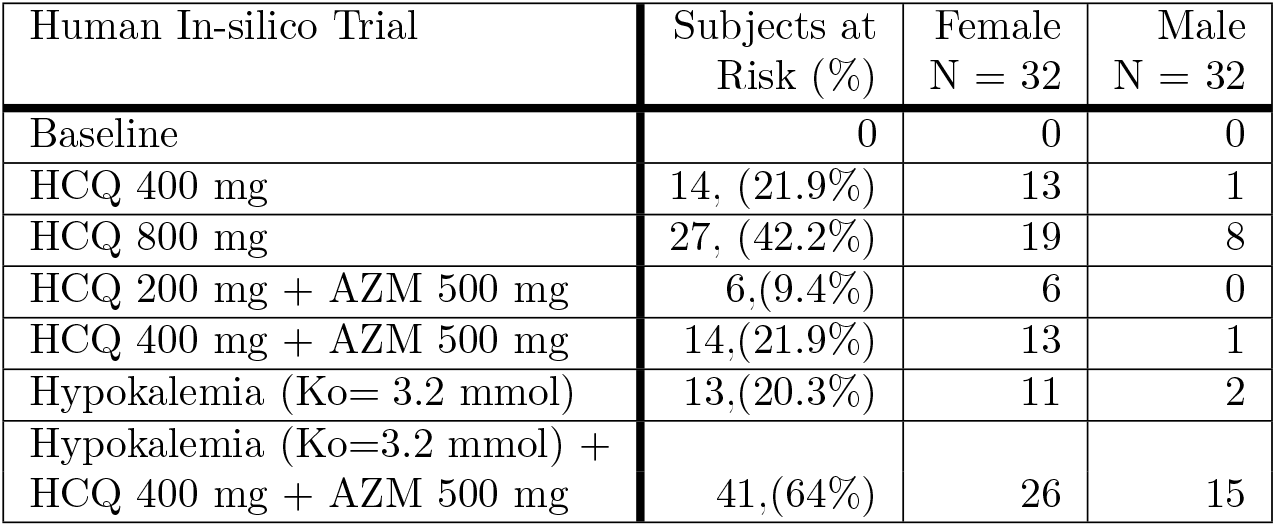
Classification based on the stress test (BCL = 400 ms) on the virtual clinical trial.

| Human In-silico Trial | Subjects at Risk (%) | Female N = 32 | Male N = 32 |
| --- | --- | --- | --- |
| Baseline | 0 | 0 | 0 |
| HCQ 400 mg | 14, (21.9%) | 13 | 1 |
| HCQ 800 mg | 27, (42.2%) | 19 | 8 |
| HCQ 200 mg + AZM 500 mg | 6,(9.4%) | 6 | 0 |
| HCQ 400 mg + AZM 500 mg | 14,(21.9%) | 13 | 1 |
| Hypokalemia (Ko= 3.2 mmol) | 13,(20.3%) | 11 | 2 |
| Hypokalemia (Ko=3.2 mmol) + HCQ 400 mg + AZM 500 mg | 41,(64%) | 26 | 15 |

**Fig 8.**
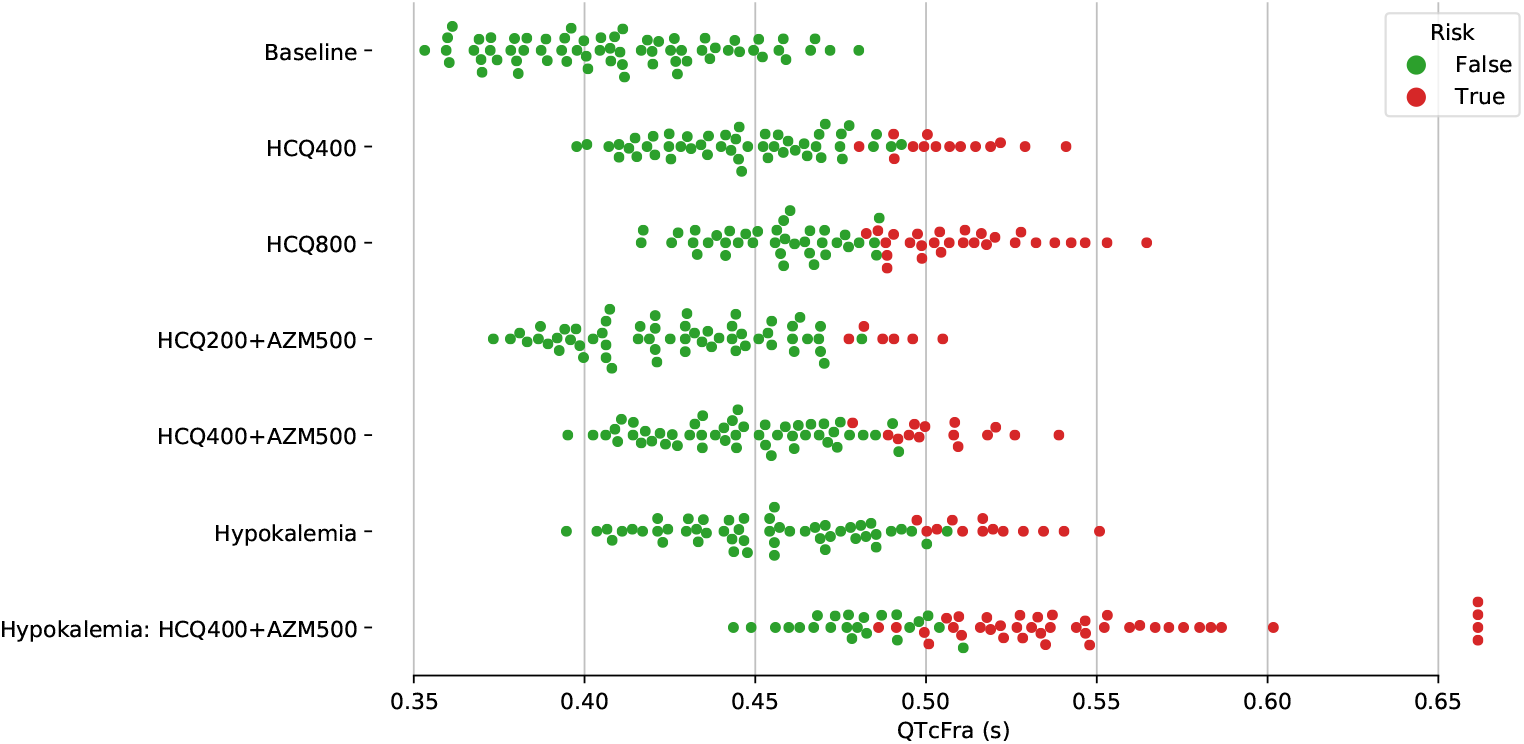
**QTcFra results of all cohorts** classified by risk. Subjects at risk of drug-induced arrhythmias are shown in red. QTcFra shown is the one calculated at the baseline BCL of 600ms.

**Fig 9.**
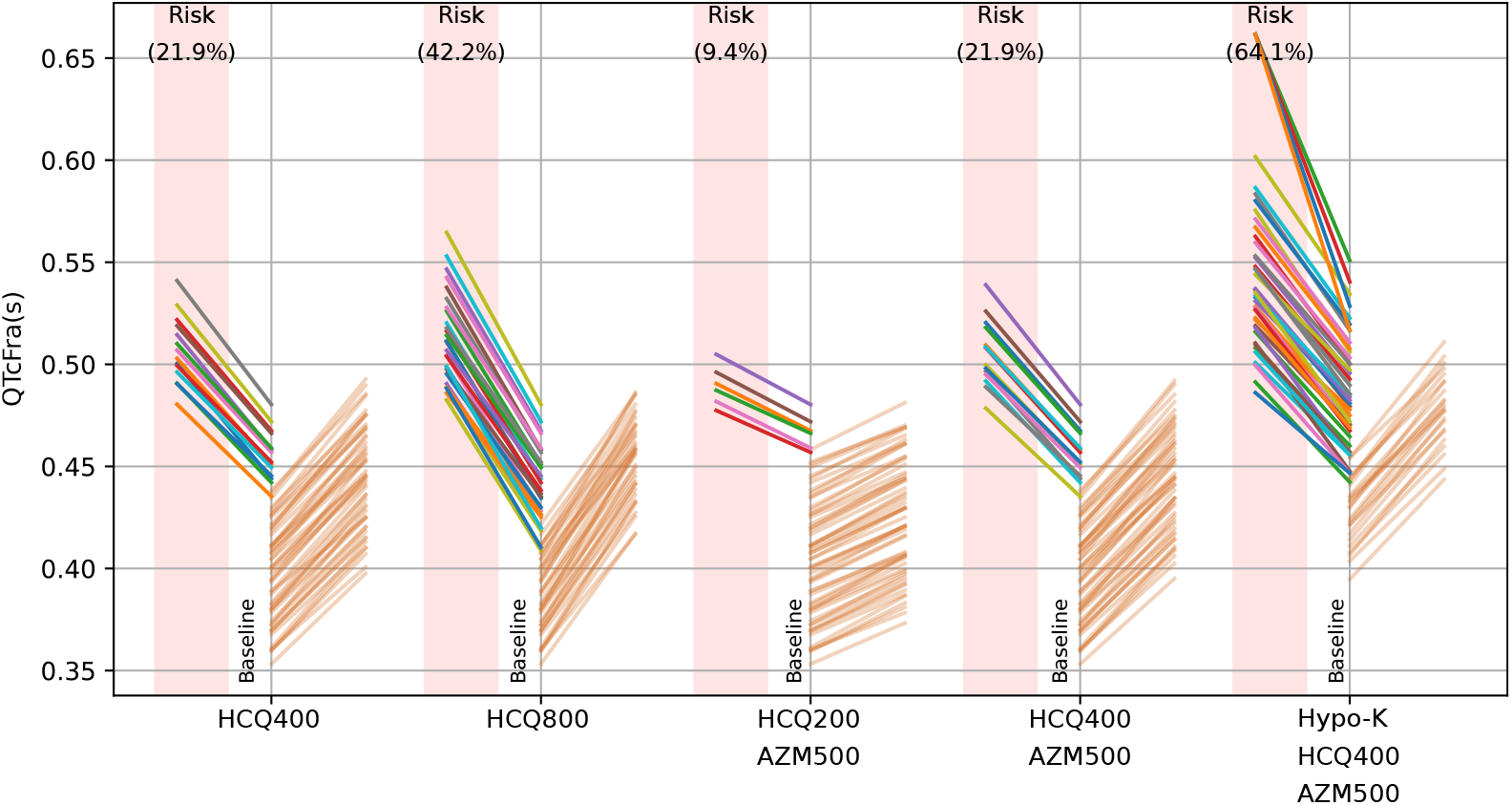
**QTcFra prolongation trajectory of each subject** of the normal and hypokalemic virtual population after the administration of various doses of HCQ (oral dosis in mg) as sole drug, and in combination with AZM (oral dosis in mg) classified by risk. Each line connects the values of QTcFra at baseline and treated, on the left are the lines corresponding to the subjects at risk, while on the right are the remaining subjects. Note the overlap of the lines on the baseline axis, demonstrating poor performance of QTc as risk indicator.The QTcFra corresponds to the baseline BCL of 600ms.

The characteristics of the arrhythmic phenotypes were traced back within the baseline population, in order to identify the phenotypes that might exhibit a propensity for a drug-induced arrhythmic behavior. The most common phenotype in the virtual population at high risk, was a low GKr, however not all of the phenotypes with low GKr elicited these arrhythmic scenarios (see Fig. 10). The relative importance of these currents show that the main current involved in the QTc length was GKr; however, the relative importance for the QTc prolongation after the administrations of the various drug doses, were GKs and GNaL.

**Fig 10.**
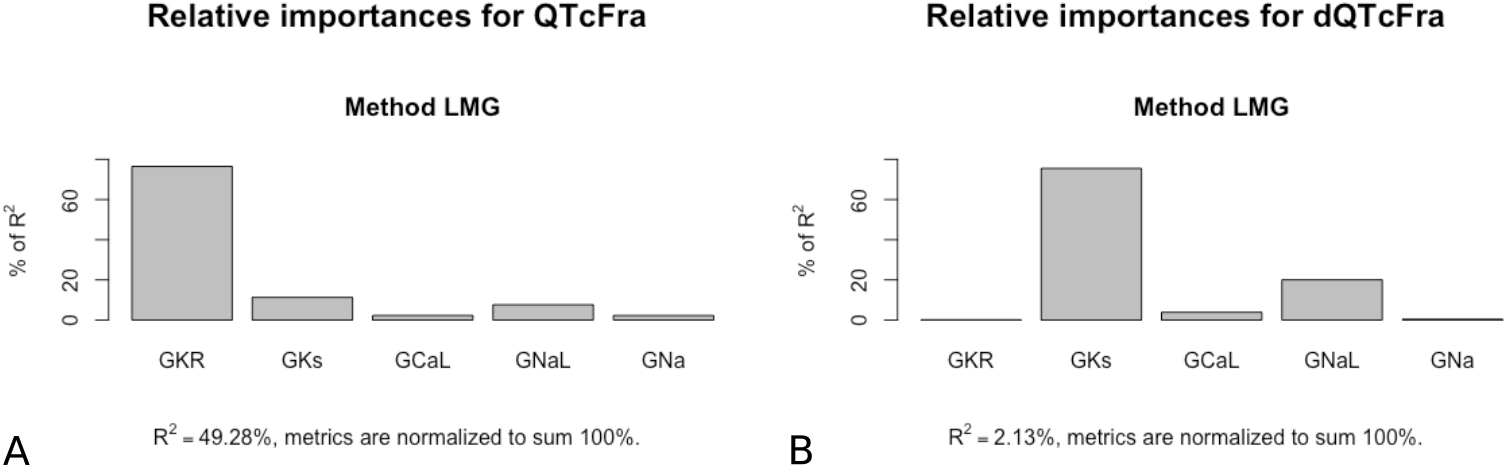
**LMG relative importance of the ion channel current conductances** on two of the measured markers: A) QTcFra and B) the prolongation of QTcFra.

Within this study, QTc was normalized employing both the Framingham (QTcFra) and Bazett (QTcBaz) convention. It is noticeable that both normalizations provide different QTc values. The Bazett convention provides QTc values consistently higher than the Framingham convention at 600 ms BCL (on average by 52.5 ± 15.9 ms higher).

### In-vitro experiments

The recorded in-vitro measurements from the reanimated swine hearts are shown in Table 6. Pressure-volume loops of the left ventricle at baseline and after the administration of various doses of HCQ are shown in Fig. 11. The entire protocol is shown in different plots in order to observe the full dataset that overlaps after the first, washout and second dose. Two main aspects should be pointed out from the haemodynamic response of the in-vitro setup to large doses of HCQ: the hemodynamic function was detrimentally affected and it did not recover after washout of the drug.

**Table 6.** In-vitro functional response to the administration of 7.44 mol/lt of HCQ and after full drug washout. (mean± std).

|  | BCL<br>(s) | LVPsystolic<br>(mmHg) | LVPdiastolic<br>(mmHg) | dP/dt<br>(mmHg/s) | Stroke Work<br>(mL mmHg) |
| --- | --- | --- | --- | --- | --- |
| Baseline | 0.604 ± 0.002 | 76.9 ± 5.56 | 18.39 ± 0.39 | 948.17 ± 289.2 | 179.48 ± 6.81 |
| 25 mg dose 1 | 0.586 ± 0.135 | 50.3 ± 9.12 | 19.99 ± 1.3 | 396.2 ± 107.63 | 58.2 ± 27.4 |
| Washout | 0.578 ± 0.133 | 60 ± 10.97 | 16.43 ± 3.83 | 597.49 ± 107.14 | 81.45 ± 30.8 |
| 25 mg dose 2 | 0.598 ± 0.06 | 87 ± 5.73 | 30.28 ± 5.07 | 542.55 ± 125.99 | 60.8 ± 12 |
| 50 mg | 0.711 ± 0.121 | 49.14 ± 6.5 | 18.26 ± 7.5 | 375.66 ± 88.08 | 61.5 ± 23.2 |

**Fig 11.**
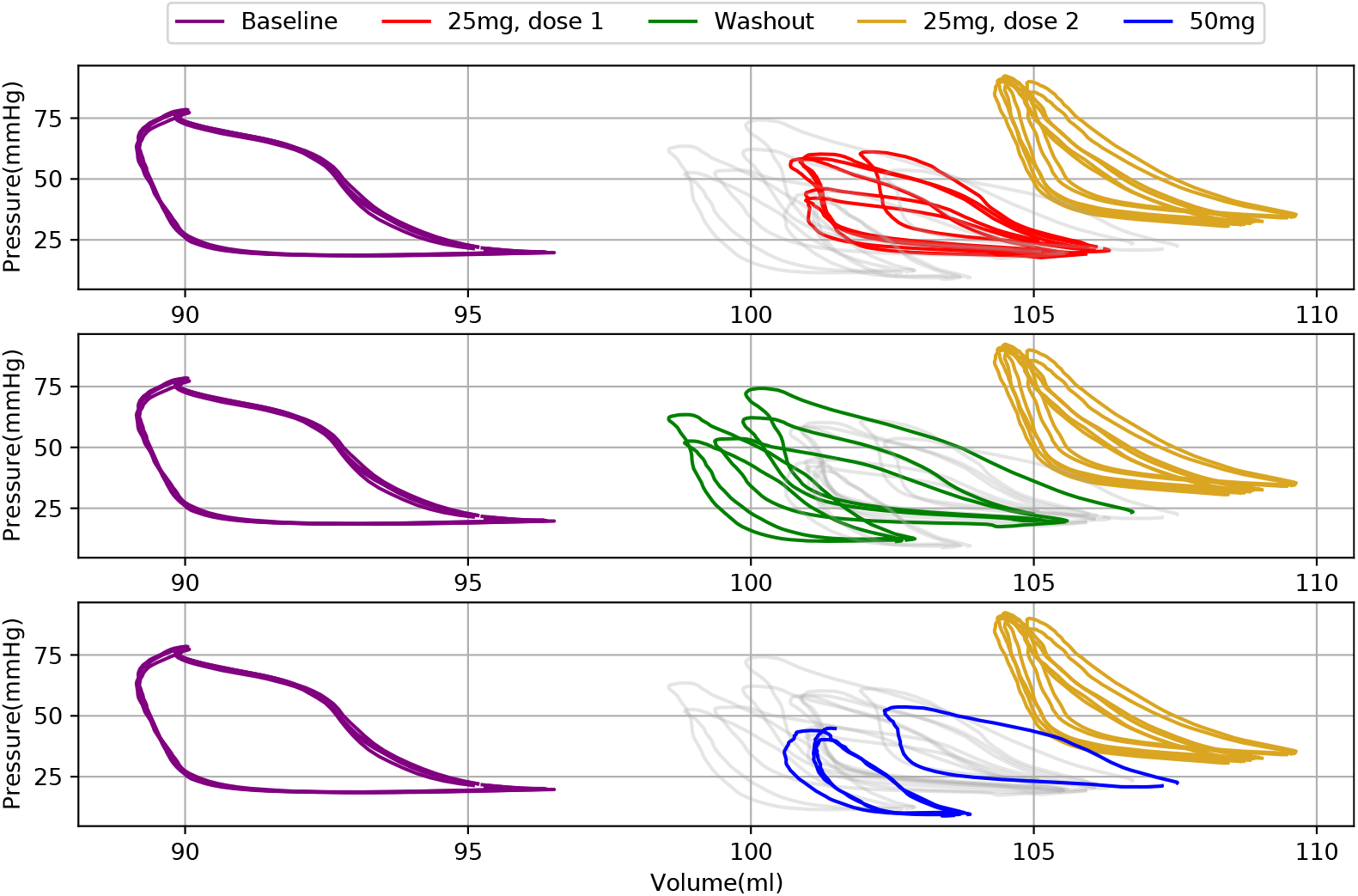
**Characteristic left ventricular pressure-volume loops** from an ex-vivo perfused, reanimated swine heart at baseline and following administration, washout, and re-administration of HCQ. The entire protocol is shown in three panels due to overlapping data (in gray color as reference). Baseline and second dose of 25 mg are included in all panels for ease of comparison. First 25 mg dose is shown in red in the top panel, washout in green in the middle panel and second 25 mg dose in blue in the third panel. Notice the variability observed in the haemodynamics after the administration of large doses of HCQ. Hemodynamic function was not recovered after washout.

Monophasic action potentials measured in the ex-vivo swine heart (Fig. 12) show the APD after the administration of a large dose of HCQ. Epicardial MAP measurements elicited an APD increase after a 25 mg administration of HCQ from 352 to 363 ms (APD prolongation of 11 ms) in the epicardium, and from 350 to 481 ms increase in the endocardium (APD prolongation of 131 ms). For the extremely high dose of 50 mg (14.88 µmol/l) the APD in the epicardium increased to 589 ms (APD prolongation of 237 ms). Interestingly, the APD differences between endocardium and epicardium on a virtual subject where LBBB was observed after the administration of 800 mg of HCQ, showed an increase of the AP in the endocardium of an average 0.063 ± 0.0315 s with respect to the epicardium. Measurements are shown in supplemental video S3.

**Fig 12.**
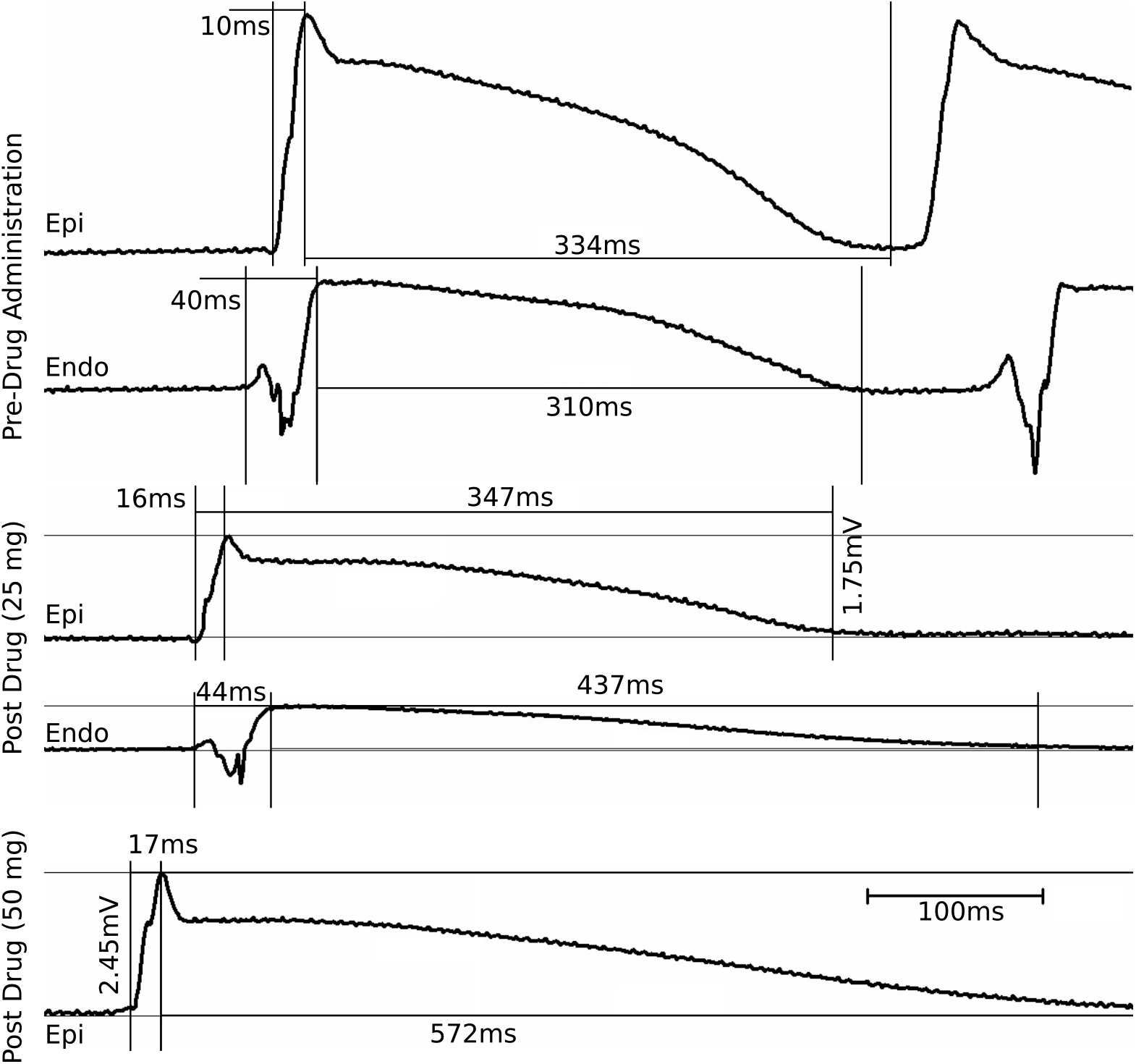
MAPs recorded from the endocardial and epicardial walls of a reanimated swine heart. Note, only epicardial MAPs were able to be recorded for the 50mg HCQ dose.

## Discussion

This is the first study of drug-induced arrhythmic risk after the administration of one or more potentially cardiotoxic drugs using a 3D full heart “virtual patient population”. The developed computational framework employs gender-specific cardiac phenotypes under both normal and hypokalemic conditions. We specifically sought to identify alterations induced by the administrations of hydroxychloroquine (HCQ) and azithromycin (AZM). A series of experiments on reanimated swine hearts to assess function and types of arrhythmic risks within an in vitro model after the administration of HCQ were also performed.

The QTc values and QTc prolongations obtained from the computational framework fell within the ranges observed on clinical patients. The results were compared to those of clinical studies recently published [45, 46]. In both clinical studies, the Bazett normalization was employed as shown in Table 7.

**Table 7.** Comparison of QTc Bazett values between the virtual clinical trial and published clinical trials.

| QTc Bazett (ms) | Virtual Clinical Trial<br>N= 64, Median [iqr], Mean ± std | Mercuro et al [46]<br>N = 90, Median [iqr] | Saleh et al [45]<br>N = 200, Mean ± std |
| --- | --- | --- | --- |
| Baseline | 447.5[413-481], 449.4 ± 41.3 | 455[430-474] | 440.6 ± 24.9 |
| HCQ 400 mg | 509[476-546], 512.8 ± 44.6 | 473[454-487] | NA |
| HCQ 200 mg + AZM 500 mg | 475[444-511], 477.9 ± 42.2 | NA | 470.4 ± 45.0 |
| HCQ 400 mg + AZM 500 mg | 508[477-543], 511.5 ± 44.2 | 442[427-461] | NA |
| Hypokalemia (Ko=3.2 mmol) +<br>HCQ 400 mg + AZM 500 mg | 587 [548-627], 598.4 ± 70 | NA | NA |

The stress test results indicated that 21.9% of the computational cohort exhibited a higher arrhythmic risk after the administrations of a sole dosis of HCQ (400 mg) and the co-administration of HCQ (400 mg) plus AZM (500 mg). For the administration of 200 mg of HCQ plus 500 mg of AZM, the percentage risk fell to 9.4%. The risk increased significantly in subjects with hypokalemia (3.2 mol K+ concentration) to 64.1% within the virtual population. At a peak plasma concentration after the administration of 800 mg of HCQ, the risk for arrhythmic events was calculated as 42.2%. The virtual clinical trial was able to confirm that the use of HCQ was associated with a proarrhythmic risk, but it did not worsen when it was used in combination with AZM as has been observed clinically, experimentally and in-silico [9, 45, 46].

It is important to stress that the concentrations tested within these virtual healthy patients were peak plasma concentrations. These predicted values may change according to the relative metabolisms of these drugs in a clinical setting and according to the interaction with other anti-arrhythmic medications. Nevertheless the proposed framework allows for the testing of any plasma concentration of a given drug on cardiac electrophysiology.

Comparison to clinical data published by Saleh et al. [45] is difficult, provided that all subjects, even with a variety of potassium concentrations lower than normal, were included within their cohort statistics. However, it is noticeable that ventricular tachycardia occurred in 7 out of 8 of the reported patients with a potassium concentration lower than normal, in agreement to the high risk shown in the virtual hypokalemic population, Table 8. The match between the percentages of subjects at risk after the administration of 400 mg of HCQ and the combination of 400 mg of HCQ and 500 mg of AZM was remarkable as compared to [46].

**Table 8.** Comparison of the percentage of subjects at risk between the virtual clinical trial and the clinical trials published in the literature.

| Subjects at Risk (%) | Virtual Clinical Trial N= 64 | Mercuro et al. [46] N = 90 | Saleh et al. [45] N = 200 |
| --- | --- | --- | --- |
| HCQ 400 mg | 21.8% | 19% | NA |
| HCQ 200 mg + AZM 500 mg | 9.3% | NA | 3.5% |
| HCQ 400 mg + AZM 500 mg | 21.8% | 21% | NA |
| Hypokalemia (K<sub>o</sub>=3.2 mmol) + HCQ 400 mg + AZM 500 mg | 64% | NA | NA |

The in-vitro experimentation on the reanimated swine hearts provided evidence for transmural heterogeneous effects of HCQ on action potential durations as shown in Fig. 12. This was evidenced by heightened drug-induced transmural heterogeneities, as has been previously associated with multi-channel blocking drugs [47]. Enhanced transmural heterogeneities of focal APs provide an important arrhythmic risk, as observed in one of the subjects that presented an LBBB after the administration of 800 mg of HCQ. This further confirms the capability of the computational models to exhibit the arrhythmic behaviors observed experimentally. Lastly, the long-lasting effects on cardiac function after a complete washout of a large dose of HCQ was an important observation towards the clinical use of HCQ. One limitation to the electrophysiology simulations in this study is the lack of an electro-mechanical simulation that could provide mechanistic information regarding the detrimental haemodynamic effects of the large dose of HCQ observed in the in-vitro study. An extension to the solution of an electro-mechanical model to assess drugs in a large population like the one proposed here, would require an approximate 6.5 times more computation time. While running electro-mechanics was not an aim of this work, it constitutes a part of our future work.

There is a limitation to this study related to the IC50 values obtained from the literature [36, 38]. To date, they have not been quantified on data obtained for human cardiomyocytes; these data come from guinea pig SAN cells and human ion channels heterologously expressed in human embryonic kidney HEK293 cells. These values, however, provided a similar response in our virtual population to recently published clinical trials. The translation of ion channel behaviors between species regarding cardiac ion channel functions remains today, an extremely complex unsolved problem. In this work, we employed the only data available in the literature at the time this study was performed. Furthermore, the interactions of two administered cardiotoxic drugs also remains a complex problem. The ‘naive’ approach assumed in this work, regarding the additive effect of the two drugs on the ion channel blocking effect, can be revised as emerging research becomes available. All effective plasma concentration and IC50 data on the drugs employed can be better adjusted within the proposed framework as they become available. Another important limitation of this work is the high computational cost of the study. Computational time availability on commercial and research HPC infrastructures, however, is increasing constantly and is not a limiting factor to insilico human heart trials. The authors are aiming to produce simulations that may aid in the reduction of human clinical trials, according to the established three Rs. Therefore, provided the adequate platform and computational time accessibility, these workflows may be integrated within the drug development pipeline. The results from the simulations reflect the behavior of a normal and hypokalemic population without any additional risk factors (i.e. ischemia, other electrolyte disturbances, infarction, and/or established cardiac genetic disorders). The assessment of the drug effects on a more broad population remains part of future work. Furthermore, the assessment of the drug induced pro-arrhythmic risk in a variety of human cardiac anatomies is also part of the future work. Nevertheless, the assumptions tested in the present study were able to provide timely critical information concerning phenotypic characteristics of the subset of our virtual subjects that have a higher predisposition to drug-induced QT prolongations. The use of more detailed ion channel models of herg can be employed in future work.

## Conclusion

A virtual gender-specific cohort yielded a range of electrocardiographic phenotypes that resemble a normal human cohort. The computational framework developed was capable of reproducing the effects of one or more administered cardiotoxic drugs, showing remarkable resemblance to published clinical studies. It was capable of reproducing the complexity of the cardiotoxicity response in the human population. Furthermore, it may be employed in future urgent applications to provide primary information about the dosages and the combined effects of drugs; most importantly this has application when clinical guidance is unavailable. The data required for such computational models are the plasma concentrations of the drug or drugs in question and the IC50 values for each of the cardiac ion channels they affect. An in-silico clinical trial framework like the one proposed in this work, could be capable of providing evidence of the proarrhythmic risk of QTc-interval prolonging drugs in a normal or diseased population using high performance computing within a day.

The virtual normal cohort can further be interrogated relative to any population variants that produce a distinct, arrhythmogenic outcome after the administration of one or various drug combinations. This can be exploited towards the identification of more accurate potential markers of cardiotoxicity. The identification of the most prevalent phenotypes within a specific population may further be employed within this computational framework: i.e, to establish predictive bounds of arrhythmogenesis of a relevant, specific sample population.

## Data Availability

Human cardiac database is hosted at the Visible Heart Laboratory, University of Minnesota website.
Computational model results will become available after peer review publication.

## Supporting information

**S1 Video**. LBBB produced in a female subject after the administration of 800 mg of HCQ.

**S2 Video**. Ventricular tachycardia observed in a female subject after the administration of 400 mg of HCQ and 500 mg of AZM.

**S3 Video**. LBBB produced in a female subject after the administration of 800 mg of HCQ including endocardial vs epicardial APs.

**S1 Table**. Median (interquartile range); mean ± standard deviations of all the markers assessed in the entire population at baseline and after the administration of 400 mg and 800 mg of HCQ; and the administration of 400 and 200 mg HCQ combined with 500 mg of AZM.

